# Exploring Prevalence and Drivers of Perimenopause Uncertainty Among U.S. Women: A Mixed-Methods Study

**DOI:** 10.64898/2025.12.24.25342960

**Authors:** Yihan Xu, Deirbhle Fergus, Yella Hewings-Martin, Carley Prentice, Adam Cunningham, Mary Hedges, Chrisandra Shufelt, Stephanie Faubion, Liudmila Zhaunova

**Affiliations:** Flo Health Inc., London, UK; Institute of Epidemiology & Health, University College London, London, UK; School of Population Health, Royal College of Surgeons in Ireland, Dublin, Ireland; Department of Medicine, Mayo Clinic, 4500 San Pablo Road, Jacksonville, Florida 32224, USA; Center for Women’s Health, Mayo Clinic, 200 1st St SW, Rochester, Minnesota 55905, USA

**Keywords:** Perimenopause, mixed-methods, content analysis, perimenopause uncertainty, symptom attribution

## Abstract

**Objective:** Perimenopause is an under-recognized life stage that may be accompanied by complex and fluctuating symptoms. We aimed to quantify the prevalence of perimenopause uncertainty and explore the underlying drivers.

**Methods:** We conducted a mixed-methods study based on a cross-sectional survey of U.S. women aged 35 years and above (N=7,640). Closed-ended responses were analyzed to estimate the prevalence of perimenopause uncertainty with subgroup differences investigated by age and symptom severity. Content analysis of free-text responses (n=409) was conducted to identify the main uncertainty drivers.

**Results:** Overall, 34% of participants reported being unsure of their reproductive stage. Uncertainty peaked among those aged 40-44 (42%) and was highest among those with severe symptom burden (37%). The content analysis revealed three main uncertainty drivers. Symptom confusion and attribution was the most common (56%), reflecting difficulties interpreting bodily changes and distinguishing perimenopause from other causes. Knowledge gaps and information seeking accounted for 28% of responses, highlighting limited health literacy, assumptions about age, and active searches for evidence. Barriers to confirmation and care (16%) described dismissive healthcare encounters and reluctance to acknowledge perimenopause. Younger women (35-39 years) were more likely to cite knowledge gaps, while healthcare barriers peaked in the 40-44 age group.

**Conclusion:** Perimenopause uncertainty is a prevalent and clinically meaningful challenge. This uncertainty is conceptually distinct from illness-focused models: it is a universal transition with ambiguity and often lack of validation. Better symptom recognition and targeted communication is a crucial first step toward improving women’s awareness and support during perimenopause.

## Introduction

Perimenopause is the transitional period leading up to the final menstrual period (FMP) and including the first 12 months of amenorrhea, typically marked by considerable hormonal changes ^1,2^. Perimenopause usually begins in the mid-40s, though onset can vary widely, with a median age of onset around 45 years and a median duration of roughly 4-8 years. ^3,4^. In the United States (U.S.) alone, approximately two million women are estimated to begin perimenopause each year ^5^. This period can meaningfully affect women’s wellbeing, with more than half (59% - 65%) experiencing vasomotor, psychological, or urogenital symptoms ^6^, which have been shown to impair daily functioning, diminish quality of life, and reduce work productivity ^7–9^.

Despite the fact that it is a common life stage, recognizing perimenopause remains a challenge for both individuals and clinicians. The Stages of Reproductive Aging Workshop +10 (STRAW+10) framework defines typical cycle patterns, and to a lesser extent, hormonal changes and symptom profiles characteristic of perimenopause ^10^. However, no single biomarker reliably confirms perimenopause onset. Even standard markers such as follicle-stimulating hormone (FSH) fluctuate widely both between reproductive stages and within the same day such that they offer limited diagnostic clarity ^1,11^.

Meanwhile, perimenopause symptom profiles are heterogeneous and may evolve throughout ^12,13^. These symptoms also overlap with a wide range of other conditions, including premenstrual syndrome (PMS), thyroid disease, and mental health issues, making attribution difficult when cycle changes are minimal or vasomotor symptoms are not present ^14–17^. This complexity may fuel perimenopause uncertainty even in the presence of substantial symptom burden.

Limited public awareness and inconsistent clinical recognition further compound this uncertainty. Many women report little prior knowledge about perimenopause, hold assumptions based on age, or have difficulty distinguishing perimenopause from stress or other health conditions ^16,18,19^. Clinicians, meanwhile, often receive inadequate training and education on perimenopause or menopause care, contributing to dismissive or inconclusive assessments in the primary care setting ^17,20^. Together, these symptom and informational challenges create and sustain uncertainty about whether one is in perimenopause.

Perimenopause uncertainty can be partially understood through existing theoretical frameworks, though none fully captures its unique features. Mishel’s Illness Uncertainty Theory conceptualizes uncertainty as conditions arising from ambiguous symptoms, limited cues, or inconsistent information, which strongly parallels the perimenopause experience ^21^. However, perimenopause differs from the illness contexts as it is a natural life transition rather than a disease state. Transitions Theory ^22^ similarly highlights vulnerability during major life changes, but assumes transitions to be visible, acknowledged, and often medically guided – conditions that frequently do not apply to perimenopause. Han and colleagues’ ^23^ taxonomy of medical uncertainty further distinguishes uncertainty by sources (ambiguous or incomplete information, complex symptom patterns), issues (scientific questions about reproductive staging, practical decisions about care-seeking), and locus (experienced by patients, clinicians, or jointly). While conceptually valuable, the taxonomy has not been empirically applied to the perimenopause uncertainty phenomenon. Altogether, those theories underscore the need for more empirical work that moves beyond theoretical description to examine how uncertainty manifests among women navigating perimenopause.

Prior studies have described elements of perimenopause uncertainty, such as confusion about symptoms, misconceptions about age, and invalidating healthcare encounters, but most are small, focus on women above 40, and are rarely U.S.-based ^16,19,24–26^. Crucially, no study has hitherto quantified how common perimenopause uncertainty is, what its common drivers are, and whether those drivers differ across age groups or levels of symptom burden. Moreover, mixed-methods approaches remain rare, despite their ability to pair population-level estimates with lived accounts ^27^.

In this study, we sought to address evidence gaps by analyzing data from a large cross-sectional survey of U.S. women aged 35 and above who took part in a survey about perimenopause. Using a mixed-methods approach, we aimed: (1) to quantify the proportion of women who report feeling uncertain about their perimenopause stage and investigate how it varies by age and symptom burden; (2) to explore the underlying reasons for this uncertainty through qualitative analysis of open-text responses; and (3) to examine whether the key drivers of uncertainty vary by age group and symptom burden.

## Methods

### Study design and ethics

A convergent mixed-methods design was used to capture both the prevalence and the underlying reasons for uncertainty about perimenopause. A mixed-methods approach was chosen because, while several standalone qualitative studies have explored women’s experiences of perimenopause, none to our knowledge have combined quantitative and qualitative evidence to examine uncertainty at scale. With a large dataset of over 7,600 U.S. women aged 35 and above, integration of closed- and open-ended responses allowed quantification of the magnitude of uncertainty and to complement these findings with women’s own explanations, thereby providing a more comprehensive account than either method alone ^28^.

We obtained ethical approval for the survey and analyses from the WCG Institutional Review Board (IRB No. 20244604). All participants provided informed consent prior to beginning the survey, and responses were collected anonymously via an online survey platform, SurveyMonkey.

Standards for Reporting Qualitative Research (SRQR) was followed ^29^ and the Good Reporting of A Mixed Methods Study (GRAMMS) guidelines ^30^ were used in reporting this study.

### Study participants and data collection

Data was collected between December 6th, 2024, and May 16th, 2025, as part of a large survey about attitudes toward and symptom experiences during perimenopause. In total, 13,122 U.S. based participants visited the survey page (Figure 1). Participants who did not complete the survey (n = 4,685) or were currently using hormonal birth control (n = 797) were excluded from analysis. This resulted in 7,640 participants.

**Figure 1.**
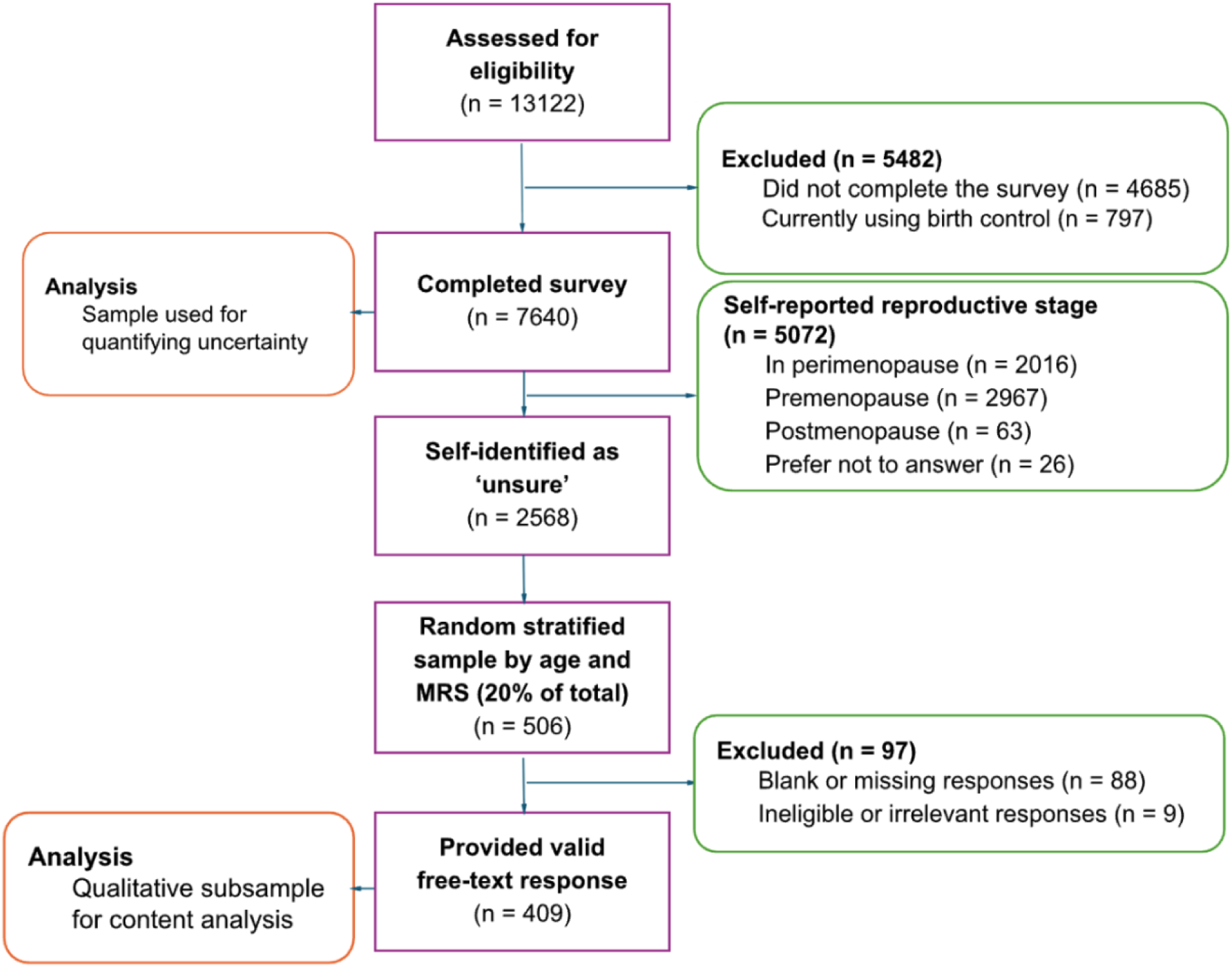
Participants flow chart. The process of determining eligibility for study inclusion for both quantitative and qualitative components of the study.

All participants were presented with standardized definitions of reproductive stage (premenopause, perimenopause, postmenopause) and asked to select the stage that best described them (see Supplementary material A). Of those eligible, 5,072 self-reported a reproductive stage, while 2,568 selected “I am unsure.” This group is referred to as the unsure analytic subsample. Although the survey item asked about several reproductive stages, qualitative responses indicated that women overwhelmingly interpreted ‘unsure’ as uncertainty specifically about whether they were in perimenopause. For clarity and consistency with participant interpretations, this construct is thereafter referred to as perimenopause uncertainty.

For the quantitative data collection, beyond self-reported reproductive stage, we collected symptom severity as measured by the Menopause Rating Scale (MRS) ^31^ and demographic characteristics.

For the qualitative data collection, participants who identified as unsure were offered the option to provide a free-text explanation for their uncertainty with a prompt stating: “You’ve told us that you’re unsure, would you like to tell us why?” A random stratified subsample of 20% of unsure participants was obtained (n = 506), stratified by age (35-39, 40-44, 45-49, 50+) and by MRS severity levels (minimal: 0-4; mild: 5-8; moderate: 9-15; severe: 16+). Stratification ensured that the subsample captured a balanced range of symptom severity and age groups, while also keeping the volume of qualitative data manageable for in-depth manual coding. Of the subsample, 409 participants provided valid free-text responses, forming the qualitative subsample for content analysis.

### Data analysis

#### Quantitative strand

We analyzed closed-ended survey responses to estimate the prevalence of perimenopause uncertainty and examined subgroup differences. Frequency analysis was utilized to assess the dispersion of categories across the data, and Chi Square tests at 5% significance levels were employed to assess whether categories within subgroups significantly differed. For the purposes of calculating intercoder reliability (ICR) and frequency analysis, each unit of analysis was assigned the most prominent code. All quantitative analyses were conducted in R (version 4.4.1).

#### Qualitative strand

Conventional content analysis ^32^ was used to analyze the free-text responses data. This method is well suited for analyzing large-scale survey data, as it can combine inductive coding with frequency analysis ^33^. Unlike thematic analysis, which is often applied to smaller, interview-based samples, content analysis is epistemologically compatible with mixed-methods designs, as it accommodates both descriptive (code frequencies, category counts) and interpretive (theme development) elements ^34,35^, allowing identification of common explanations for uncertainty, while also quantifying their relative prominence across subgroups. An inductive approach was taken when coding as it is applicable to under-explored topics like perimenopause as it is most suited to surface explanatory categories not pre-specified by researchers and to inform targets for education and care during perimenopause ^34^.

The unit of analysis was each individual free-text response [Table 1]. Two coders were involved in the analysis. An initial set of 40 codes was generated through open coding in Microsoft Excel. These preliminary codes were reviewed for conceptual overlap and redundancy, and thematically related or overly narrow codes were then combined and consolidated, resulting in a coding frame of 25 codes, each accompanied by a coding list documenting inclusion/exclusion criteria and examples [Supplementary Table S1]. ICR was assessed through independent application of the codebook to 10% of the data, yielding an overall Cohen’s Kappa of .85, indicating strong agreement. All discrepancies were resolved through discussion. Codes were grouped into categories and subcategories by relevance to the research questions, enabling both frequency analysis and interpretation of explanatory themes.

**Table 1.** Example from the process of content analysis.

| Unit of analysis | Code | Sub-category | Category |
| --- | --- | --- | --- |
| *My periods are still regular, but I have symptoms like hot flushes, weight gain, mood changes* | Symptom-cycle conflict | Change to cycles and symptoms | Symptom<br>Confusion & Attribution |

#### Rigor and reflexivity

The Standards for Reporting Qualitative Research ^36^ were followed to ensure transparency in both the research and reporting process. Principles of credibility, dependability, confirmability, and transferability were followed to ensure trustworthiness of the findings ^37^. The analysis was conducted by two researchers with expertise in women’s health and qualitative methods. Artificial intelligence assistance in content analysis was considered ^38^, but was decided against due to data security and ethical concerns.

## Results

### Participants characteristics

Across the full U.S. sample aged 35 years and above (n = 7,640), the mean age was 41.3 years (SD = 5.0). Participants were ethnically diverse with varied levels of education and most identifying as White (63%). Symptom burden was high with about two-thirds reported moderate or severe menopause symptom burden by MRS scores. The unsure analytic subsample (n = 2,568) was broadly similar to the full sample, though it included more participants aged 40-44 years, had lower educational attainment, and showed higher symptom severity. The qualitative subsample (n = 409) closely mirrored the full unsure group across key characteristics, consistent with being a stratified random sample [Table 2].

**Table 2.**
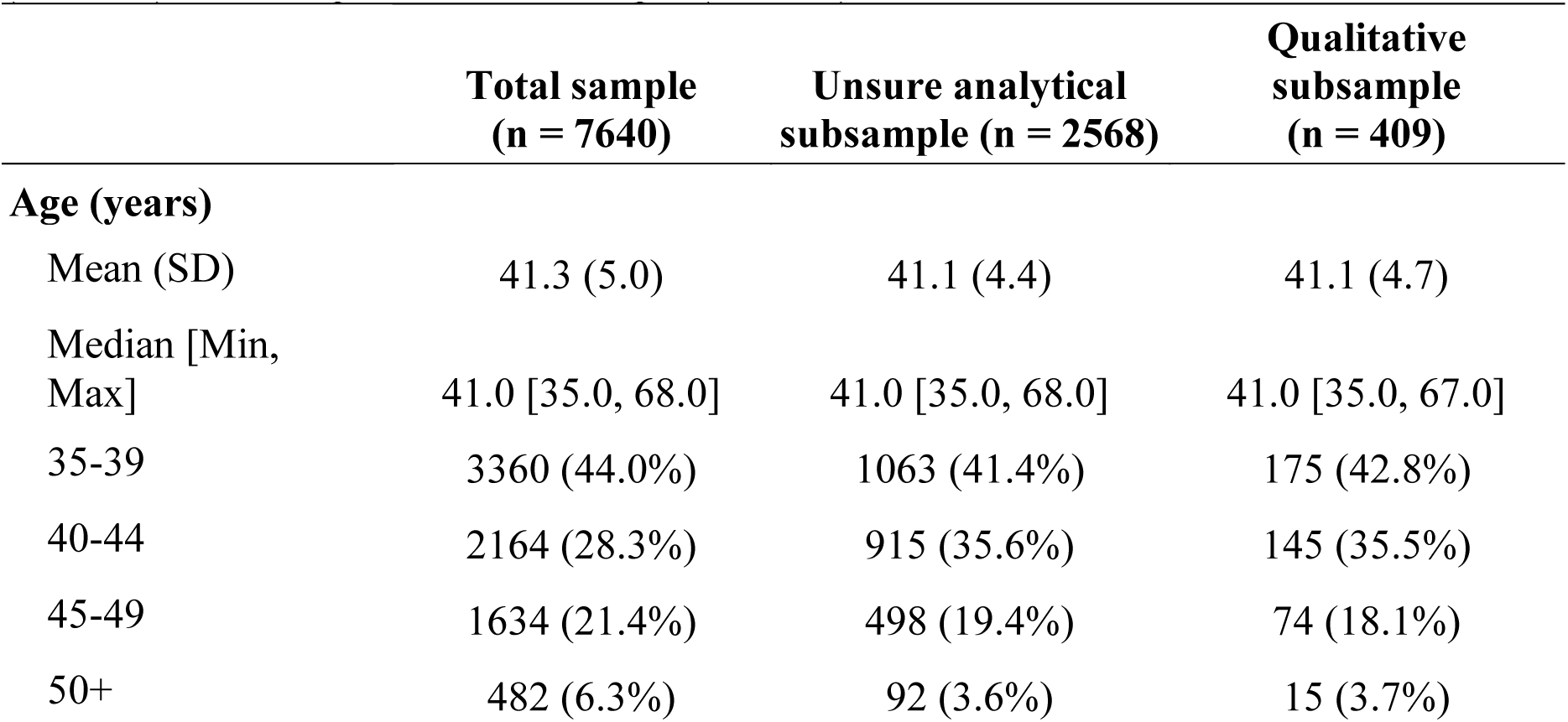
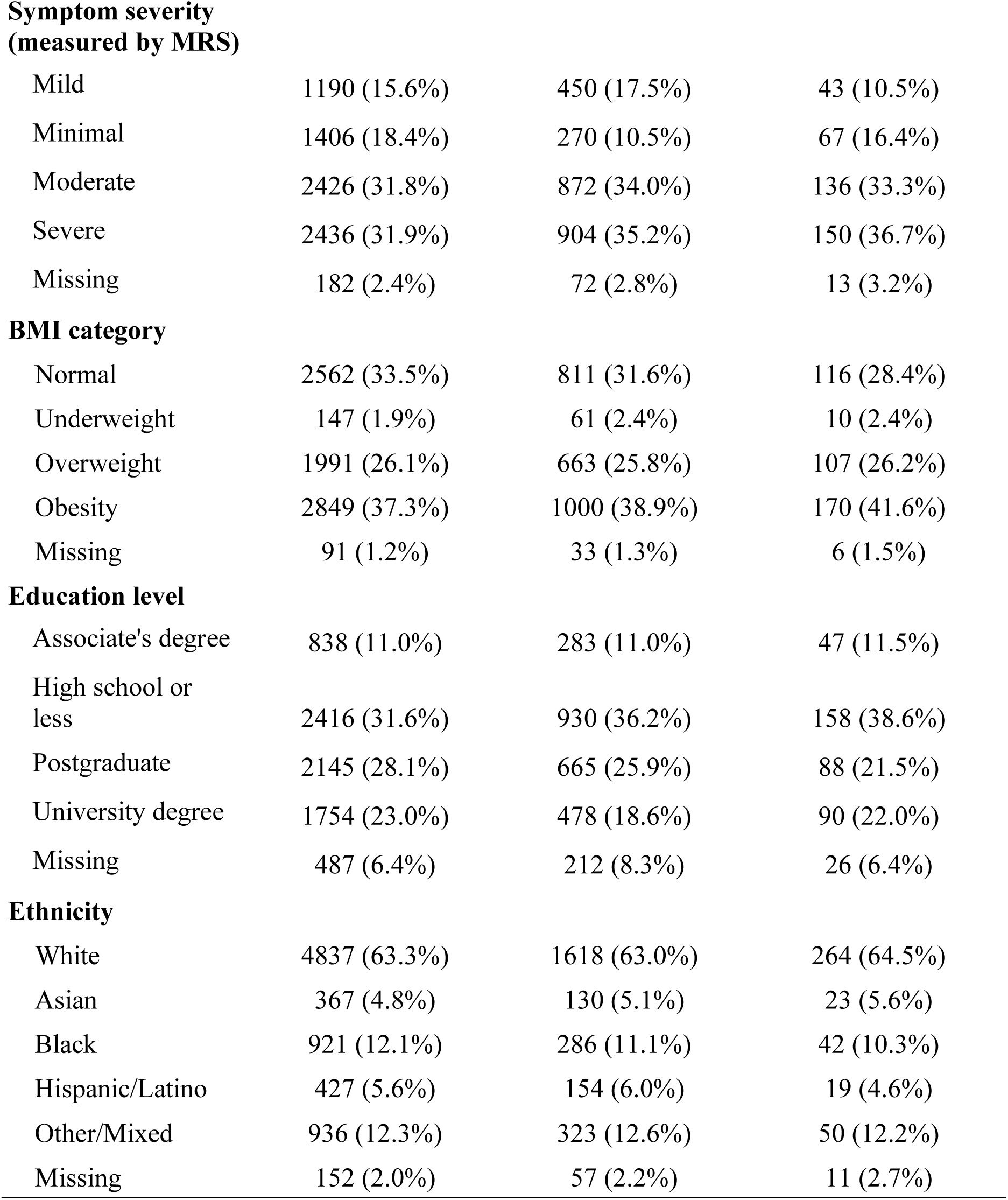
Participant characteristics of total sample (n=7,640), unsure analytic subsample (n=2,568), and the qualitative subsample (n = 409).

|  | Total sample<br>(n = 7640) | Unsure analytical<br>subsample (n = 2568) | Qualitative<br>subsample<br>(n = 409) |
| --- | --- | --- | --- |
| **Age (years)** |  |  |  |
| Mean (SD) | 41.3 (5.0) | 41.1 (4.4) | 41.1 (4.7) |
| Median [Min,<br>Max] | 41.0 [35.0, 68.0] | 41.0 [35.0, 68.0] | 41.0 [35.0, 67.0] |
| 35-39 | 3360 (44.0%) | 1063 (41.4%) | 175 (42.8%) |
| 40-44 | 2164 (28.3%) | 915 (35.6%) | 145 (35.5%) |
| 45-49 | 1634 (21.4%) | 498 (19.4%) | 74 (18.1%) |
| 50+ | 482 (6.3%) | 92 (3.6%) | 15 (3.7%) |
| **Symptom severity (measured by MRS)** |  |  |  |
| Mild | 1190 (15.6%) | 450 (17.5%) | 43 (10.5%) |
| Minimal | 1406 (18.4%) | 270 (10.5%) | 67 (16.4%) |
| Moderate | 2426 (31.8%) | 872 (34.0%) | 136 (33.3%) |
| Severe | 2436 (31.9%) | 904 (35.2%) | 150 (36.7%) |
| Missing | 182 (2.4%) | 72 (2.8%) | 13 (3.2%) |
| **BMI category** |  |  |  |
| Normal | 2562 (33.5%) | 811 (31.6%) | 116 (28.4%) |
| Underweight | 147 (1.9%) | 61 (2.4%) | 10 (2.4%) |
| Overweight | 1991 (26.1%) | 663 (25.8%) | 107 (26.2%) |
| Obesity | 2849 (37.3%) | 1000 (38.9%) | 170 (41.6%) |
| Missing | 91 (1.2%) | 33 (1.3%) | 6 (1.5%) |
| **Education level** |  |  |  |
| Associate's degree | 838 (11.0%) | 283 (11.0%) | 47 (11.5%) |
| High school or less | 2416 (31.6%) | 930 (36.2%) | 158 (38.6%) |
| Postgraduate | 2145 (28.1%) | 665 (25.9%) | 88 (21.5%) |
| University degree | 1754 (23.0%) | 478 (18.6%) | 90 (22.0%) |
| Missing | 487 (6.4%) | 212 (8.3%) | 26 (6.4%) |
| **Ethnicity** |  |  |  |
| White | 4837 (63.3%) | 1618 (63.0%) | 264 (64.5%) |
| Asian | 367 (4.8%) | 130 (5.1%) | 23 (5.6%) |
| Black | 921 (12.1%) | 286 (11.1%) | 42 (10.3%) |
| Hispanic/Latino | 427 (5.6%) | 154 (6.0%) | 19 (4.6%) |
| Other/Mixed | 936 (12.3%) | 323 (12.6%) | 50 (12.2%) |
| Missing | 152 (2.0%) | 57 (2.2%) | 11 (2.7%) |

### Prevalence of uncertainty and subgroup differences

Uncertainty about perimenopause was common with one third of women (34%, n = 2,568) reporting that they were unsure of their reproductive stage. Uncertainty varied significantly by age (χ² (3, 7,640) = 131.6, *p* <.001), peaking at over 40% among women aged 40-44, the age range when perimenopausal changes often begin, and falling to below 20% among women aged 50 and above [Figure 2, Supplementary Material A].

**Figure 2.**
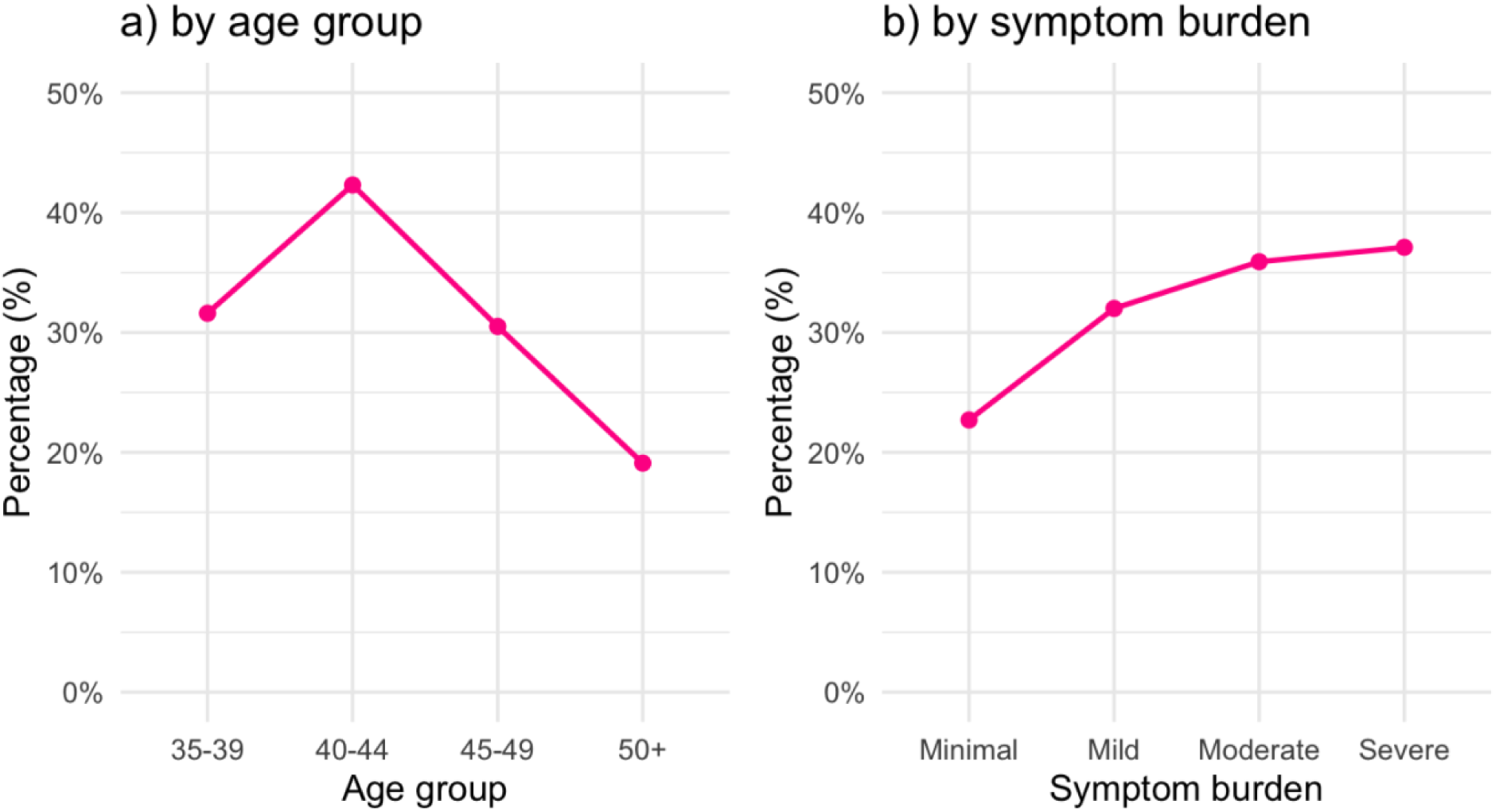
Percentage of women who reported uncertainty about their reproductive stage, stratified by (a) age group and (b) symptom burden as measured by the Menopause Rating Scale (MRS).

Overall, the likelihood of being unsure increased with symptom severity (χ² (4, 7,640) = 87.41, p < .001; Figure 2b). While fewer than one in four women with minimal symptoms reported uncertainty (23%), this rose to more than one in three among those with moderate (35%) or severe (37%) symptoms [Figure 2].

### Drivers of uncertainty

Content analysis of the 409 valid free-text responses revealed three main drivers of uncertainty: (1) Symptom confusion and attribution, (2) Knowledge gaps and information seeking, and (3) Barriers to confirmation and care. Symptom confusion and attribution was the most common, accounting for over half of responses, while the other categories captured different ways that uncertainty was reinforced through information deficits and difficulties accessing or accepting medical confirmation [Figure 3, Supplementary Figure S1].

**Figure 3.**
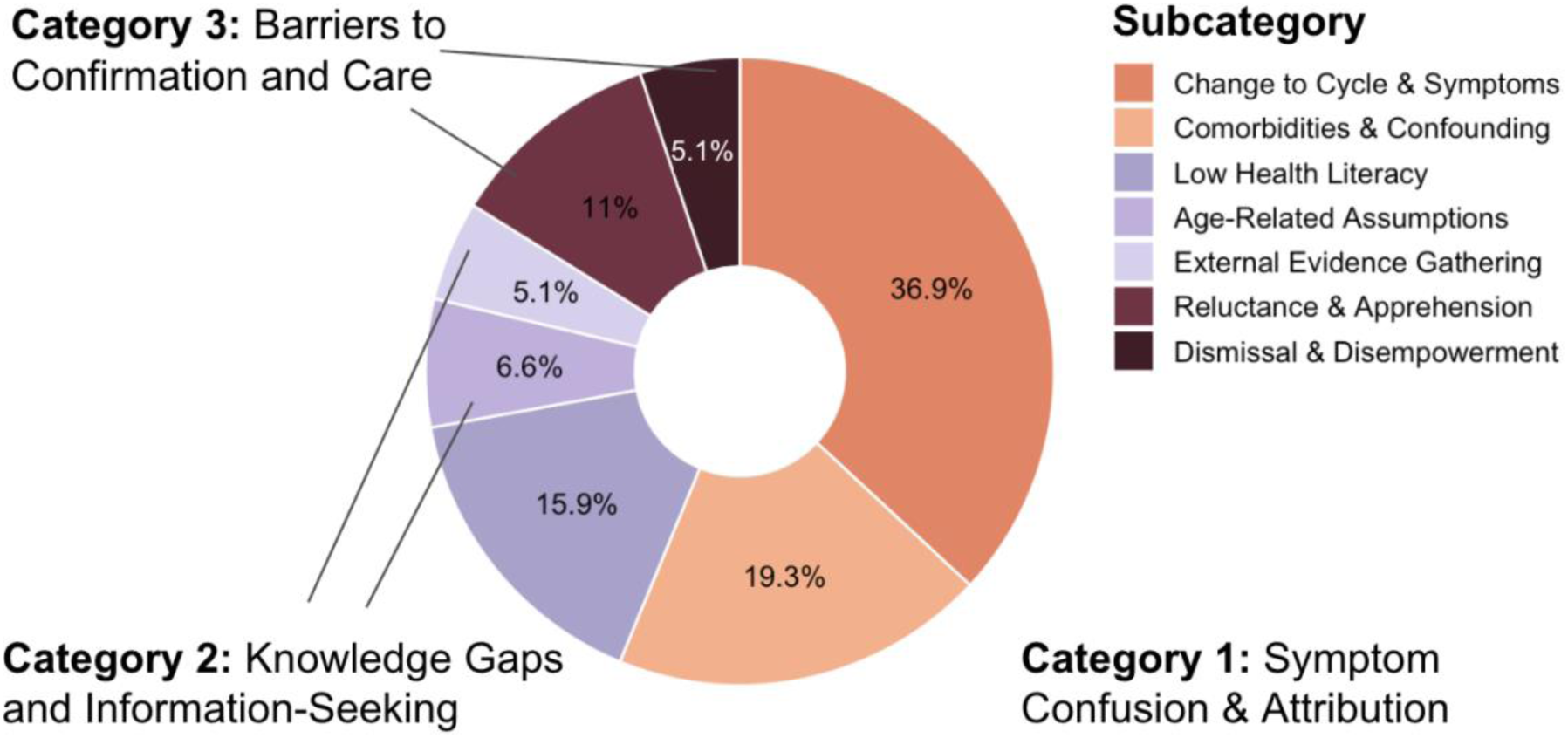
Categories and subcategories of drivers that explain perimenopause uncertainty. The most common category (56%) related to symptom confusion & attribution (orange), particularly changes to cycles/symptoms and comorbidities. Knowledge gaps & information seeking (purple) accounted for 28%, while barriers to confirmation & care (brown) represented 16%.

### Category 1: Symptom Confusion & Attribution

The most common explanation for uncertainty was symptom confusion and attribution (56% of responses, n = 230). This category included the difficulty of interpreting bodily changes and reconciling them with expectations of perimenopause. Participants often described being unsure whether their symptoms counted as perimenopause, especially when menstrual cycles remained regular or when they had other health conditions.

#### Change to cycles and symptoms

Over a third of participants linked their uncertainty to presumed menstrual cycle changes (37%, n=151). While changes to the menstrual cycle were recognized as a core indicator of perimenopause among respondents, applying this knowledge to their own individual circumstances proved difficult. For some, cycles remained regular despite experiencing classic symptoms:

- *“My periods are still regular, but I have symptoms like hot flushes, weight gain, mood changes.”* (Age: 40-44; MRS: moderate)

For others, long-standing irregular cycles made it more challenging to distinguish perimenopause-related changes from their usual pattern:

- *“My period is very irregular (always has been) and I only get it 3 ∼ 4 times a year.”* (Age: 45-49; MRS: severe)

#### Comorbidities and confounding factors

Nearly one in five women described other health conditions that blurred the boundaries between perimenopause and alternative explanations (19%, n=79). These included reproductive disorders such as endometriosis:

- *“I have irregular periods, I also have endometriosis so I don’t know if I’m in it or not.”* (Age: 40-44; MRS: moderate)

As well as hormonal conditions like thyroid disease:

- “*My symptoms may be related to hypothyroidism that I’m working to get under control.”* (Age: 40-44; MRS: minimal)

and other medical co-morbidities:

- “*I have a lot of symptoms that correspond to perimenopause, but I’m also not sure if it’s related to my anxiety, my high blood pressure, or a combination of it all.”* (Age: 40-44; MRS: severe)

Some participants highlighted other life events, such as postpartum weaning or discontinuing hormonal contraception, that can involve low-estrogen states or withdrawal effects that might come with cycle irregularity, mood swings, or vasomotor symptoms that overlap with the early perimenopause experience:

- *“I am weaning my 14-month-old so it’s hard to say if the hormone changes are due to that or are age-related.” (Age: 35-39, MRS: moderate)*
- *“I came off the contraception injection a year ago. Now my periods are back all over the place.” (Age: 40-44, MRS*: moderate)

### Category 2: Knowledge Gaps and Information-Seeking

The second most common driver of uncertainty was knowledge gaps and information-seeking (28%, n = 113). Participants expressed a medley of confusion, misunderstanding, and misconceptions surrounding perimenopause, prompting some to actively gather information. The perceptions and behaviors highlighted that uncertainty was not simply about symptoms, but also about how much they knew, or sought to know, about perimenopause.

#### Health literacy

Around one in six participants (16%, N = 59) either explicitly expressed or implicitly suggested that they did not know what perimenopause was, how it manifested, or how to recognize it in themselves. There was a clear gradient in knowledge, ranging from a complete lack of understanding of perimenopause to the perpetuation of common health myths.

Some acknowledged a fundamental knowledge gap:

- *“I don’t know the meaning of perimenopause.” (Age: 35-39; MRS: moderate)*
- *“I don’t know the symptoms.”* (Age: 40-44; MRS: severe)

Others confused perimenopause with postmenopause:

- *“I still have hot flashes, but haven’t had a period in over 10 years*” (Age: 50+; MRS: severe)

A few referenced misinformation or misattribution:

- “*I’ve started to get tremendous brain fog. Maybe it’s a covid vaccine thing. It started happening after the vaccine.*” (aged 40-44, MRS: mild)

#### Age-related assumptions

Some participants based their uncertainty on age-related assumptions about perimenopause (7%, n=27). Several women equated perimenopause with ageing and therefore discounted their symptoms as unrelated. Younger participants in particular expressed disbelief that they could already be entering perimenopause:

- *“My period has changed so much in the past 12 months and I have perimenopause symptoms but I think I’m too young.” (Age: 35-39, MRS: severe)*
- *“Too young.”* (Age: 45-49, MRS: minimal)

By contrast, a smaller group resisted the notion that age alone should determine their reproductive stage, challenging assumptions that midlife automatically meant perimenopause:

- *“Just because I’m in my 40s doesn’t necessarily mean I’m perimenopausal.*” (Age: 40-44, MRS: minimal)

#### External evidence gathering

A smaller group (5%, n=21) described deliberate efforts to seek clarity by compiling evidence from multiple sources:

- *“I’ve received conflicting information about how perimenopause is diagnosed. I’ve been told that I have to have no period for 6 months before I can be considered perimenopausal, I’ve been told I’m simply too young at age 44 for peri to even be considered, and I’ve also been told that both of those stances are incorrect and perimenopause can present differently for different individuals.”* (Age: 40-44, MRS: severe)

Some compared experiences with family history:

- “*All the women on my mom’s side underwent menopause before reaching 45.”* (Age: 40-44, MRS: moderate)

Some relied on peer conversations or social knowledge:

- ‘‘*[I] heard it usually starts later and signs are often dismissed by doctors.*” (Age: 35-39, MRS: severe)

While the hearsay was not always reliable, it did prompt some women to start to consider perimenopause:

- “*People keep saying I might be in perimenopause.*” (Age: 40-44, MRS: mild)

### Category 3: Barriers to Confirmation and Care

While less common, barriers to confirmation and care represented an important driver of uncertainty (16%, n=66). This category identified difficulties in obtaining or accepting medical confirmation of and care for perimenopause. Medical help-seeking was not straightforward for many participants. For some, uncertainty was reinforced by dismissive or contradictory healthcare encounters; for others, it reflected their own reluctance to acknowledge the transition.

#### Reluctance and apprehension

A sizable minority (11%, n=45) of participants described their uncertainty as rooted in their own hesitancy to acknowledge perimenopause. For some, particularly among the younger women under 40, avoidance stemmed from emotional resistance:

> *“I suspect I might be [in perimenopause], I guess I’m resistant to the idea that that’s the stage I’m in.”* (Age: 35-39, MRS: moderate)

For others, it was tied to fear of change or reluctance to confront what perimenopause symbolized:

> *“I’m 6 days late to my period and it has never happened before. I’m not pregnant, so I fear this might be the beginning of perimenopause.”* (Age: 40-44, MRS: mild)

Taking help-seeking action regarding perimenopause fundamentally relies on acceptance, and for many participants, there was a deference to their healthcare clinician for confirmation or diagnosis:

> *“I do not have an official diagnosis, but experience a lot of related symptoms. I am still waiting for answers.”* (Age 35-39, MRS: severe)

#### Dismissal and disempowerment

Some women (5%, n=21) did attempt to engage with healthcare clinicians to confirm if they were in perimenopause but felt their concerns were minimized or dismissed, often on the grounds of age.

- “*I have all the symptoms, but my healthcare provider keeps telling me I am too young.*” (Age: 40-44, MRS: severe)

…or symptom profile:

- “*When I saw my OB/GYN last year she told me I didn’t have enough symptoms.*” (Age: 45-49, MRS: moderate)

Others reported receiving alternative diagnoses, such as depression, or conflicting advice between their clinicians:

- *“The doctor told me it was depression.” (Age: 35-39, MRS: severe)*
- *“[I] recently saw a consultant rheumatologist [who] suspects evidence of [perimenopause] symptoms… GP insists [it’s] not perimenopause.”* (Age: 45-49, MRS: severe)

These encounters left some participants feeling frustrated, unheard, or invalidated:

- *“Doctors just perform a pap and run labs and say I’m normal, but I know my body, and I say different.”* (Age: 40-44, MRS: missing)

### Subgroup difference of uncertainty drivers

To explore whether the underlying reasons for perimenopause uncertainty varied across demographic and health profiles, we examined the distribution of qualitative categories by age group and symptom severity [Figure 4]. While all three categories appeared across subgroups, we observed notable differences in their relative prominence by age and symptom burden.

**Figure 4.**
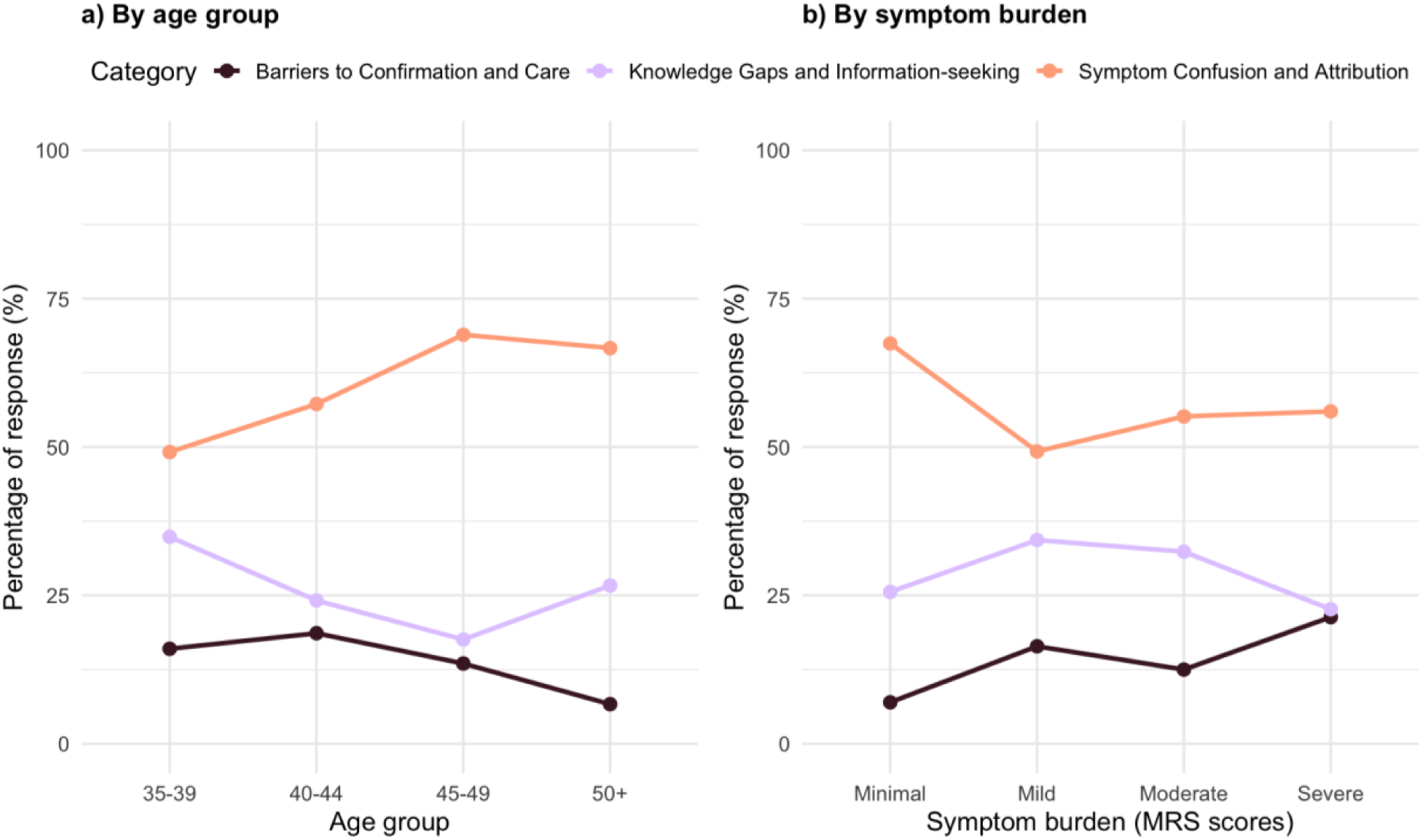
Drivers of uncertainty about perimenopause, by a) age group and b) symptom burden. Percentages represent the proportion of responses coded to each category within subgroups. Across all groups, symptom confusion and attribution (orange) was the most prominent driver, and it increased with age but decreased with symptom severity. Knowledge gaps and information-seeking (purple) was least mentioned in the 45-49 age group, whereas more often mentioned among women with mild to moderate symptoms. Barriers to confirmation and care were most evident among younger women and those with severe symptoms (brown).

#### Age-related differences of uncertainty drivers

Differences across age groups were modest but significant (χ²(6) = 12.37, p = 0.05). Symptom confusion and attribution dominated in every group but peaked among women aged 45-49 (69%). Knowledge gaps and information-seeking were more frequent among younger women aged 35-39 (35%). Conversely, barriers to confirmation and care were most frequent in the 40-44 group (19%). Among women aged 50 and above, symptom confusion and attribution remained high (67%), but barriers to confirmation and care became less common, reflecting that clinicians may be more ready to recognize perimenopause or menopause at this stage. However, knowledge gaps and information-seeking persisted (27%).

#### Differences by symptom burden of uncertainty drivers

Variation across symptom-severity groups was non-significant (χ²(8) = 13.67, p = 0.09), but several patterns emerged. Symptom confusion and attribution remained consistently high across all symptom levels and peaked when symptoms were minimal (67%). In contrast, women with mild or moderate symptoms more often mentioned knowledge gaps and information-seeking. Among women with severe symptoms, barriers to confirmation and care were most pronounced (21%).

## Discussion

This mixed-methods study provides one of the first population-level estimates of perimenopause uncertainty for U.S. women aged 35 and above. Uncertainty about perimenopause was substantial with one in three women unsure of their reproductive stage. Uncertainty peaked among women aged 40-44, the age group most likely to be approaching perimenopause, given that the median age of onset is 45 years ^4^. Uncertainty was also more common among women experiencing more severe symptoms. Together, these patterns reveal a striking paradox: the women who could benefit most clearly from identifying their menopause stage (e.g., those likely to be entering perimenopause and those most affected by the symptoms), appear to be the ones where it is the hardest to recognize.

Through qualitative content analysis, we identified three drivers of perimenopause uncertainty. The most common drivers were symptom confusion and attribution, knowledge gaps and information-seeking, and barriers to confirmation and care. These uncertainty drivers are not static but appear to evolve with age and symptom severity. Younger women (aged 35-39) were more likely to experience knowledge-based uncertainty, whereas for women aged 45 and above, uncertainty was more symptom-based, coinciding with the natural trajectories of symptom burden changes during perimenopause ^13,39,40^. In addition, symptom-based uncertainty was most common when symptoms were minimal. Conversely, barriers-based uncertainty was most prevalent among women with the most severe symptoms.

### Comparison with prior work and theoretical advancement

The qualitative content analysis identified three core drivers of perimenopause uncertainty which resonate with existing literature, while the quantitative analysis quantified their relative prominence.

The most prevalent driver, symptom confusion and attribution, illustrates complexity-based uncertainty. Consistent with previous research, participants had difficulty attributing non-specific symptoms to perimenopause when cycles remained regular ^41^, and experienced confusion due to overlapping comorbidities such as polycystic ovary syndrome or endometriosis ^24,42^. This study extends these findings in two important ways. First, by quantifying the relative prominence, it demonstrates that diagnostic complexity is the most common form of uncertainty across age groups. Second, this uncertainty peaks among women aged 45-49, which coincides with the typical age range for perimenopause, when symptoms often begin or become most pronounced ^3,4^. For many participants, perimenopause functioned as a diagnosis of exclusion, with uncertainty persisting until other explanations could be ruled out.

The second driver, knowledge gaps and information-seeking, represents ambiguity-based uncertainty. Consistent with prior research showing that women often have limited reproductive health literacy ^43,44^, this study found that participants had limited understanding of what constitutes as perimenopause. Many participants believed they were too young to be experiencing perimenopause, reflecting persistent stereotypes about the typical age of onset ^42^, even though symptoms may start as early as the mid-thirties ^45^. In line with Opayemi ^46^, we also found that women’s attempts to resolve their uncertainty through information-seeking often heightened confusion due to questionable source quality or conflicting information. By quantifying its prominence, this study identifies ambiguity-based uncertainty as a commonly shared driver of uncertainty. This type of uncertainty was most common among women aged 35-39 and those with mild or moderate symptoms.

The third driver, barriers to confirmation and care, reflects shared uncertainty between women and healthcare professionals and encompasses both internal and external barriers. Internally, similar to earlier research documenting fear and ambivalence during the climacteric period ^47,48^, some participants expressed apprehension to acknowledge perimenopause, often associating it with aging or loss of fertility. These attitudes may discourage help-seeking and reinforce uncertainty. Externally, participants often described being dismissed by clinicians who attributed their symptoms to stress or age, particularly when their cycles remained regular. This mirrors earlier findings that women with more subtle menstrual changes are frequently disregarded in clinical settings ^49^. Although less prevalent than other drivers, these barriers were most common among women aged 40-44 and those experiencing severe symptoms.

Identifying and quantifying these three drivers provides a robust empirical foundation necessary for advancing the theoretical understanding of perimenopause uncertainty. While previous studies have described women’s confusion about perimenopause or menopause symptoms or uncertainty about reproductive stage ^18,42,50,51^, this phenomenon has yet to be conceptualized as a distinctive type of health-related uncertainty.

In health contexts, illness-related uncertainty is a core psychological stressor ^52^. For example, diagnostic uncertainty has been shown to increase anxiety and distress ^53^ and reduce quality of life ^54^. Uncertainty also plays a key role in shaping health-seeking behaviors, while active coping strategies might motivate an individual to seek care, avoidant strategies may lead to feeling overwhelmed and contribute to the delay of care ^55^.

Our findings suggest that perimenopause uncertainty is distinctive from established models of medical uncertainty. While Mishel’s ^21^ Uncertainty in Illness Theory focuses on managing an illness, perimenopause is a natural and universal transition. The uncertainty is not about whether a disease is present, but when perimenopause begins and how to interpret its diffuse and variable presentations. Similarly, Meleis’ ^22^ Transitions Theory, while useful for general life transitions, often assumes a clear triggering event and acknowledged, medically-guided phases. Perimenopause, however, often lacks an unambiguous trigger and is frequently not validated by clinicians (barriers to confirmation and care), challenging these key assumptions. While Han et al.’s taxonomy ^23^ provides a useful framework of medical uncertainty (Source: complexity/ambiguity; Locus: patient/shared), the key distinction for perimenopause is persistence over several years. Unlike many medical uncertainties resolvable by definitive tests, perimenopause confirmation in clinical practice is less straightforward, and medical guidelines are scant for women under 45 years of age who have perimenopause symptoms ^45,56,57^. Therefore, perimenopause uncertainty seems inherent to the stage itself.

Based on its unique manifestation as an interpretive state of biological transition, perimenopause uncertainty presents as the subjective and protracted state experienced by midlife women that something has changed, accompanied by the inability to confidently determine or interpret one’s own reproductive stage in the setting of complex and fluctuating bodily signals, knowledge gaps regarding this natural course, and barriers to personal or clinical validation.

### Strengths and limitations

This study is unique in both quantifying and conceptualizing perimenopause uncertainty, using a large and ethnically-diverse U.S. sample that includes women aged 35 and above – an age group often excluded from previous research ^18,19,42^. By capturing perspectives from women in their mid-thirties onwards, the study broadens understanding of perimenopause uncertainty experiences.

The mixed-method approach allowed both quantification of uncertainty and qualitative exploration of its underlying drivers. While inductive content analysis has limits in interpretive depth, it was well-suited to the study’s exploratory aims and may inform future theoretical models of the transition through menopause. The integrated mixed-methods approach revealed both who was uncertain and why that uncertainty arose.

Several limitations should be noted. As recruitment occurred through the Flo app, participants may represent a more digitally engaged and health-motivated subset of users than the general population. The fully digital recruitment strategy may also have underrepresented women less likely to use mobile health applications. While the inductive approach to content analysis has been previously critiqued for its subjectivity ^58^, it was appropriate in this setting given the exploratory nature of this study and the current lack of perimenopause-specific theoretical frameworks. The qualitative subsample (n = 409) was limited due to the use of a stratified random sample to manage manual coding workload; however, codebook saturation was achieved.

### Implications for research, clinical practice, and digital health innovations

This is the first study to seek to quantify perimenopause uncertainty and identify factors which may contribute to difficulties recognizing perimenopause for women. Future research could extend this framework in various directions. Cross-cultural studies could test whether the three key drivers identified in this study are consistent across sociocultural contexts and healthcare systems. Prospective studies could examine the impact of uncertainty on health intentions, help-seeking behaviors, and clinical outcomes. Longitudinal and intervention studies could evaluate whether targeted strategies (e.g. structured education, clinician communication training, or digital self-assessment) can reduce perimenopause uncertainty and improve women’s wellbeing and engagement with care.

For clinical practice, symptom confusion and attribution, the dominant driver of uncertainty, underscores the need for clearer communication about the variability of perimenopause presentations and trajectories. Rather than over-relying on menstrual irregularity as the principal indicator based on current guidelines ^10^, clinicians and educational resources should be more open and flexible to consider the multidimensional symptom profiles of perimenopause and normalize cognitive, emotional, and physical changes that can occur earlier. Although cycle irregularity is characteristic of perimenopause ^10^, up to 25% women may not experience significant cycle changes during perimenopause ^14^. This reframing may help women integrate diverse experiences into a coherent explanatory model, thereby reducing perimenopause uncertainty and unnecessary distress.

For digital-health innovations, perimenopause uncertainty also presents an opportunity. Uncertainty in perimenopause is temporal and digital tools that provide longitudinal symptom tracking and digital self-assessment tools, such as the emerging Perimenopause Symptom Scale ^59^ and MenoScale ^60^, can help women visualize symptom burden patterns, explore symptom relief options, and decide when to consult clinicians. These tools can also help clinicians contextualize patient reports over time, bridging informational and experiential gaps.

## Conclusion

This study quantifies and conceptualizes perimenopause uncertainty, an experience affecting one in three women aged 35 and over and peaking, paradoxically, among those aged 40-44 and those with severe symptoms. Three key drivers were identified: symptom confusion and attribution (complexity), knowledge gaps (ambiguity), and barriers to confirmation and care (shared uncertainty).

The findings advance health uncertainty theory by demonstrating that perimenopause uncertainty is conceptually distinct from illness-based models. Rather that arising from pathology, it emerges from a natural, universal life transition in which uncertainty is inherent to the process itself. This shifts the focus of intervention away from cure-seeking and towards supporting women in managing an ongoing, normative developmental transition. Recognizing perimenopause uncertainty as a measurable, multidimensional construct may therefore be a crucial first step toward helping women navigate care more effectively during perimenopause.

## Data Availability

Aggregated data analyses and deidentified individual-level data may be shared upon reasonable request for the purpose of replication, secondary analysis, or further methodological development, pending permission from Flo Health UK Inc.

## Financial Disclosures / Conflicts of Interest

**YX** is a current employee of Flo Health Ltd and holds equity interests in Flo Health Ltd.

**D.F.** contributed to this research as part of a student placement program between University College London and Flo Health Inc.

**YHM** is a current employee of Flo Health Ltd and holds equity interests in Flo Health Ltd.

**AC** is a current employee of Flo Health Ltd and holds equity interests in Flo Health Ltd.

**CP** is a consultant of Flo Health Ltd.

**LZ** is a current consultant of Flo Health Ltd and holds equity interests in Flo Health Ltd.

**CLS** is an advisor for Bayer Pharmaceutics.

**SSF** is a consultant for Era Women’s Health Platform, delivers CME lectures for PriMed, AiCME, MedAll, and Medscape, and serves on the scientific advisory board for Weight Watchers.

For the remaining authors none were declared.

## Abbreviations

BMI: Body Mass Index
FMP: Final Menstrual Period
FSH: Follicle-Stimulating Hormone
GP: General Practitioner
GRAMMS: Good Reporting of A Mixed Methods Study
HCP: Healthcare Professional
ICR: Intercoder Reliability
IRB: Institutional Review Board
IUD: Intrauterine Device
MRS: Menopause Rating Scale
OB/GYN: Obstetrician-Gynecologist
PCOS: Polycystic Ovary Syndrome
PMDD: Premenstrual Dysphoric Disorder
PMS: Premenstrual Syndrome
SD: Standard Deviation
STRAW+10: Stages of Reproductive Aging Workshop +10
SRQR: Standards for Reporting Qualitative Research

## Supplementary materials

### Supplementary material A. Self-reported reproductive stage question

Below is the self-report question used to classify reproductive stage. Participants were presented with a standardized definition of premenopause, perimenopause, and postmenopause (with visual aid), and asked to select the stage that best described their current experience. This approach facilitated consistent classification based on cycle changes and symptom presentation, particularly among those without a confirmed menopause date.

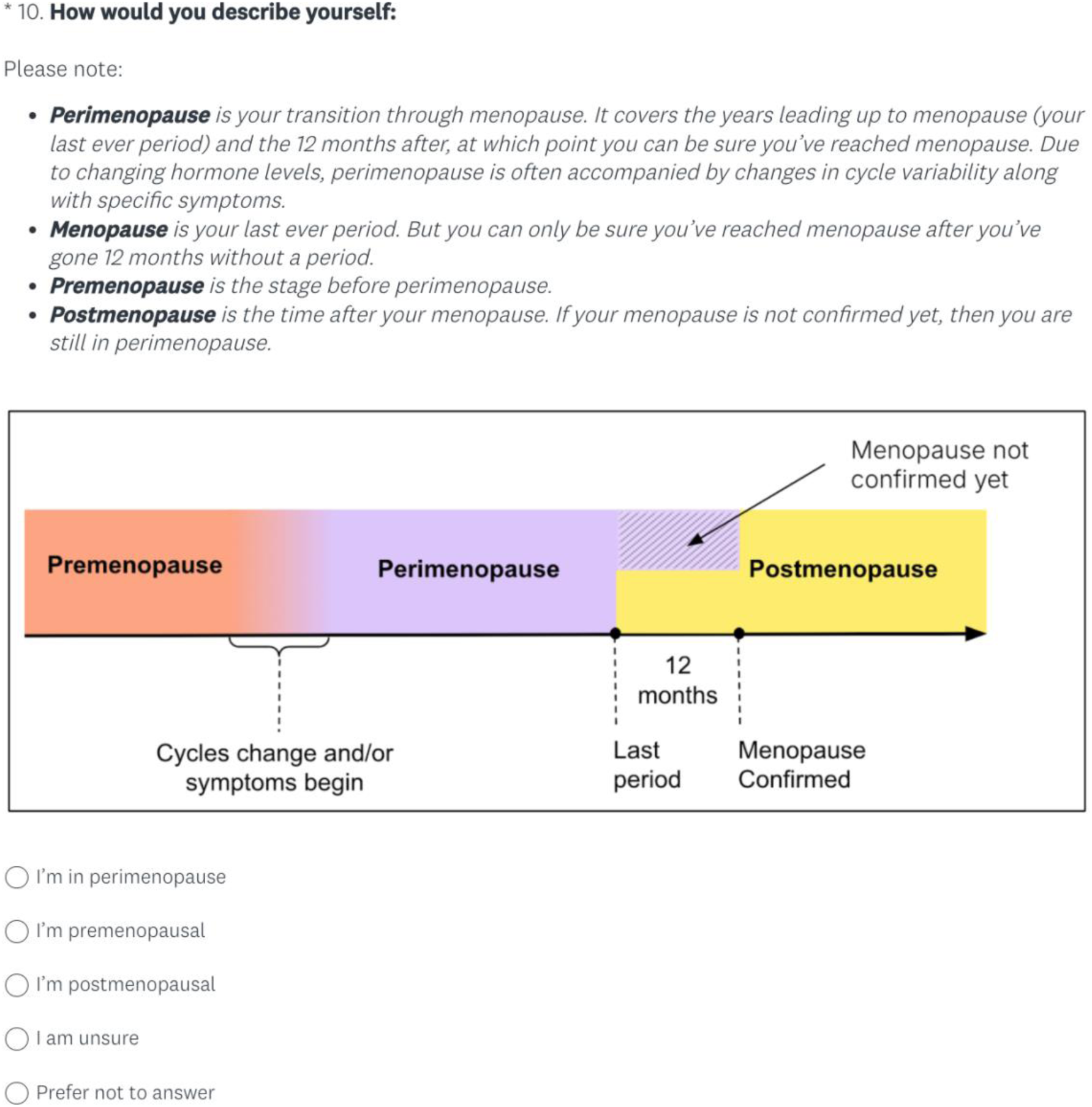

**Supplementary Table S1.** Content analysis codebook with code definition, inclusion/exclusion criteria and illustrative examples.

| Code | Inclusion/exclusion criteria | Example |
| --- | --- | --- |
| 1. Evidence gathering | Actively or passively seeking more information regarding perimenopause and symptoms | *All the research I have read seems to indicate that at my age, I must be in perimenopause but I haven't experienced any symptoms at all.* |
| 2. Hormones | May be used as evidence for or against perimenopause, or a source of confusion surrounding disentangling symptoms | *My doctor says it's just hormones.* |
| 3. Intuition | Some participants articulate 'knowing' or 'feeling' like they are in perimenopause, even in the absence of formal confirmation | *I think I am heading towards perimenopause but may not be in it yet.* |
| 4. Normal cycles | Pertains to individual descriptions of their normal cycle, cycle length, symptoms, flow and perceived 'regularity' | *I am still having a monthly menstrual cycle.* |
| 5. Lack of symptoms | The absence of symptoms of perimenopause | *I don't have all the symptoms.* |
| 6. Changes to cycle | Any notable changes to cycle length, frequency, flow, symptoms or regularity. If cycles are described as normal/regular but with some feature change, they are classified as changes to cycle rather than 'normal' cycle | *Unsure if I am perimenopausal due to still getting my period, however symptoms are nowhere as strong as when I was younger (20's).* |
| 7. Symptom recognition | Identification of perimenopause symptoms; symptoms experienced that indicated perimenopause to participant. Includes physiological and mood symptoms | *Sometimes I have the symptoms of perimenopause. My sexual drive is lower and I feel very emotional.* |
| 8. Struggling to identify symptoms | Difficulty in recognizing or attributing symptoms to perimenopause. In the case that a comorbid condition is mentioned, use comorbidities as primary code | *I feel like I am old enough to be perimenopausal but there's so much confusion around it that I have no idea if some of my symptoms are perimenopause.* |
| 9. Symptom-cycle conflict | Dissonance between cycle and symptom experience. May be perimenopause symptoms without cycle irregularity, or cycle irregularity with no additional accompanying symptoms | *I have some symptoms but still [have a] normal period.* |
| 10. Family history | Family history often served as a reference point for perimenopause; mothers', aunts' or sisters' age of menopause. Absence of family information regarding perimenopause resulted in further confusion | *All the women on my mom's side underwent menopause before reaching 45.* |
| 11. Low health literacy & misinformation | Not having enough information or knowledge to decipher symptoms or perimenopause itself. Also encompasses proliferation of incorrect 'mis' information | *I don't know what perimenopause is.* |
| 12. Age-related assumptions | Despite experiencing symptoms, describe being 'too young' or not the right age to be at perimenopause | *I'm not sure if I am at the age to experience menopause.* |
| 13. Clinician dismissal | Symptoms dismissed by doctors on the basis of age, symptom profile or cycle. Often intermingled with outdated information provided by HCPs | *I have certain symptoms that align with perimenopause but doctors don't take it seriously it seems.* |
| 14. Comorbidities (conditions) | Morbid conditions which may cause similar symptoms to perimenopause, or make distinguishing symptoms challenging. This includes physical and mental health conditions ranging from endometriosis to depression | *I'm not sure if my PMDD symptoms are getting worse or if it's perimenopause.* |
| 15. Surgery | When surgery is mentioned as contributing to uncertainty surrounding menopause, due to cycle disruption or symptoms implicated in recovery. In the case that both a comorbid condition and surgery are mentioned, surgery is a secondary code | *I had a partial hysterectomy approximately ten years ago.* |
| 16. Medication | Some medications may cause side effects which mask or mirror perimenopause symptoms | *I've been on zoladex for 6 months. I'm not sure about my period cycle or symptoms as yet.* |
| 17. (in)fertility & pregnancies | Includes difficulties conceiving, infertility, ability to conceive, current or recent pregnancy, childbearing, miscarriage and breastfeeding; all of which may be used as indicators for or against perimenopause—or may contribute to confusion surrounding symptom attribution | *I thought I might be but then got pregnant but then recently miscarried so it's hard to tell.* |
| 18. Birth control | Includes references to birth control, such as oral contraceptives or IUD. Being on birth control may mask symptoms of perimenopause, or the side effects of stopping birth control may cause symptoms such as cycle disruption | *I was on birth control for 16 years and only been off birth control for almost a year and a half. My menstrual still hasn't gotten back to normal with a regular flow.* |
| 19. Perimenopause apprehension | Anxiety around impending perimenopause or surrounding attributing symptoms to perimenopause | *Currently, I'm 6 days late to my period and it has never happened before. I'm not pregnant, so I fear this might be the beginning of perimenopause, but can't be sure.* |
| 20. Stress | Stress at times was attributed as a potential cause of perimenopause symptoms | *Seems like I'm in perimenopause, but I'm in such a stressful situation (war zone), that stress might be the reason for my symptoms.* |
| 21. Awaiting confirmation | Awaiting diagnostic clarity from an authority (HCPs) but not actively seeking it. Passive approach | *I don't know if I'm perimenopausal or not. My doctor hasn't said I am either.* |
| 22. Seeking confirmation & help-seeking | Participants describe seeking out confirmation and certainty from an authority; most frequently doctors or medical testing | *I have some symptoms but blood tests say I am not in perimenopause.* |
| 23. Self-monitoring | Tentatively monitoring perimenopause symptoms, demonstrating vigilance. Often precursor to external help-seeking | *I think I'm starting to enter perimenopause based on the changes I'm observing, but not sure yet* |
| 24. Others' opinions | Other peoples' perspectives or opinions regarding perimenopause status. Used when source of information or opinion is not disclosed, or is informal | *People keep saying I might be in perimenopause.* |
| 25. Feeling of helplessness | Inability to proceed or treat symptoms as reliant on medical confirmation, or unclear what steps can be taken | *I don't know if I have any symptoms indicating I started premenopause. I wish there were ways I could know.* |

**Supplementary Table S2.** Standards for Reporting Qualitative Research (SRQR) Checklist.

| **Item number** | **Item name** | **Guidance for reporting** | **Location** |
| --- | --- | --- | --- |
| **Report section: Title and abstract** |  |  |  |
| 1 | Title | Concise description of the nature and topic of the study Identifying the study as qualitative or indicating the approach (e.g., ethnography, grounded theory) or data collection methods (e.g., interview, focus group) is recommended | Title page |
| 2 | Abstract | Summary of key elements of the study using the abstract format of the intended publication; typically includes background, purpose, methods, results, and conclusions | Abstract |
| **Report section: Introduction** |  |  |  |
| 3 | Problem formulation | Description and significance of the problem/phenomenon studied; review of relevant theory and empirical work; problem statement | Introduction |
| 4 | Purpose or research question | Purpose of the study and specific objectives or questions | Introduction - final paragraph on aims |
| **Report section: Methods** |  |  |  |
| 5 | Qualitative approach and research paradigm | Qualitative approach (e.g., ethnography, grounded theory, case study, phenomenology, narrative research) and guiding theory if appropriate; identifying the research paradigm (e.g., postpositivist, constructivist/interpretivist) is also recommended; rationale. | Methods - study design and ethics; data analysis |
| 6 | Researcher characteristics and reflexivity | Researchers' characteristics that may influence the research, including personal attributes, qualifications/experience, relationship with participants, assumptions, and/or presuppositions; potential or actual interaction between researchers' characteristics and the research questions, approach, methods, results, and/or transferability. | Methods -Rigor and reflexivity |
| 7 | Context | Setting/site and salient contextual factors; rationale. | Methods - Study participants and data collection |
| 8 | Sampling strategy | How and why research participants, documents, or events were selected; criteria for deciding when no further sampling was necessary (e.g., sampling saturation); rationale. | Methods - Study participants and data collection |
| 9 | Ethical issues pertaining to human subjects | Documentation of approval by an appropriate ethics review board and participant consent, or explanation for lack thereof; other confidentiality and data security issues. | Methods - Study design and ethics |
| 10 | Data collection methods | Types of data collected; details of data collection procedures including (as appropriate) start and stop dates of data collection and analysis, iterative process, triangulation of sources/methods, and modification of procedures in response to evolving study findings; rationale. | Methods - Study participants and data collection |
| 11 | Data collection instruments and technologies | Description of instruments (e.g., interview guides, questionnaires) and devices (e.g., audio recorders) used for data collection; if/how the instrument(s) changed over the course of the study. | Methods - Study participants and data collection |
| 12 | Units of study | Number and relevant characteristics of participants, documents, or events included in the study; level of participation (could be reported in results). | Methods - data analysis, Table 1 |
| 13 | Data processing | Methods for processing data prior to and during analysis, including transcription, data entry, data management and security, verification of data integrity, data coding, and anonymization/de-identification of excerpts. | Methods - Study participants and data collection |
| 14 | Data analysis | Process by which inferences, themes, etc., were identified and developed, including the researchers involved in data analysis; usually references a specific paradigm or approach; rationale. | Methods - Data analysis |
| 15 | Techniques to enhance trustworthiness | Techniques to enhance trustworthiness and credibility of data analysis (e.g., member checking, audit trail, triangulation); rationale. | Methods - Rigor and reflexivity |
| **Report section: Results/findings** |  |  |  |
| 16 | Synthesis and interpretation | Main findings (e.g., interpretations, inferences, and themes); might include development of a theory or model, or integration with prior research or theory. | Results - drivers of uncertainty;<br>Discussion - comparison with prior work |
| 17 | Links to empirical data | Evidence (e.g., quotes, field notes, text excerpts, photographs) to substantiate analytic findings. | Results - prevalence of uncertainty; uncertainty drivers; subgroup difference of uncertainty drivers |
| **Report section: Discussion** |  |  |  |
| 18 | Integration with prior work, implications, transferability, and contribution(s) to the field | Short summary of main findings; explanation of how findings and conclusions connect to, support, elaborate on, or challenge conclusions of earlier scholarship; discussion of scope of application/generalizability; identification of unique contribution(s) to scholarship in a discipline or field. | Discussion - principal findings; comparison with prior work |
| 19 | Limitations | Trustworthiness and limitations of findings. | Limitations and future directions |
| **Report section: Other** |  |  |  |
| 20 | Conflicts of interest | Potential sources of influence or perceived influence on study conduct and conclusions; how these were managed. | Statements and declarations<br>- Conflicts of interest |
| 21 | Funding | Sources of funding and other support; role of funders in data collection, interpretation, and reporting. | Statements and declarations<br>- funding statements |

**Supplementary Table S3.** Good Reporting of A Mixed Methods Study (GRAMMS) checklist.

| Item number | Guideline | Location |
| --- | --- | --- |
| 1 | Describe the justification for using a mixed methods approach to the research question | Introduction - second to last paragraph;<br>Methods - study design and ethics |
| 2 | Describe the design in terms of the purpose, priority and sequence of methods | Methods - data analysis |
| 3 | Describe each method in terms of sampling, data collection and analysis | Methods - data analysis: quantitative strand, qualitative strand |
| 4 | Describe where integration has occurred, how it has occurred and who has participated in it | Methods - rigor and reflexivity |
| 5 | Describe any limitation of one method associated with the present of the other method | Methods - data analysis; Discussion - limitations and future directions |
| 6 | Describe any insights gained from mixing or integrating methods | Discussion - principal findings |

**Supplementary Figure S1.**
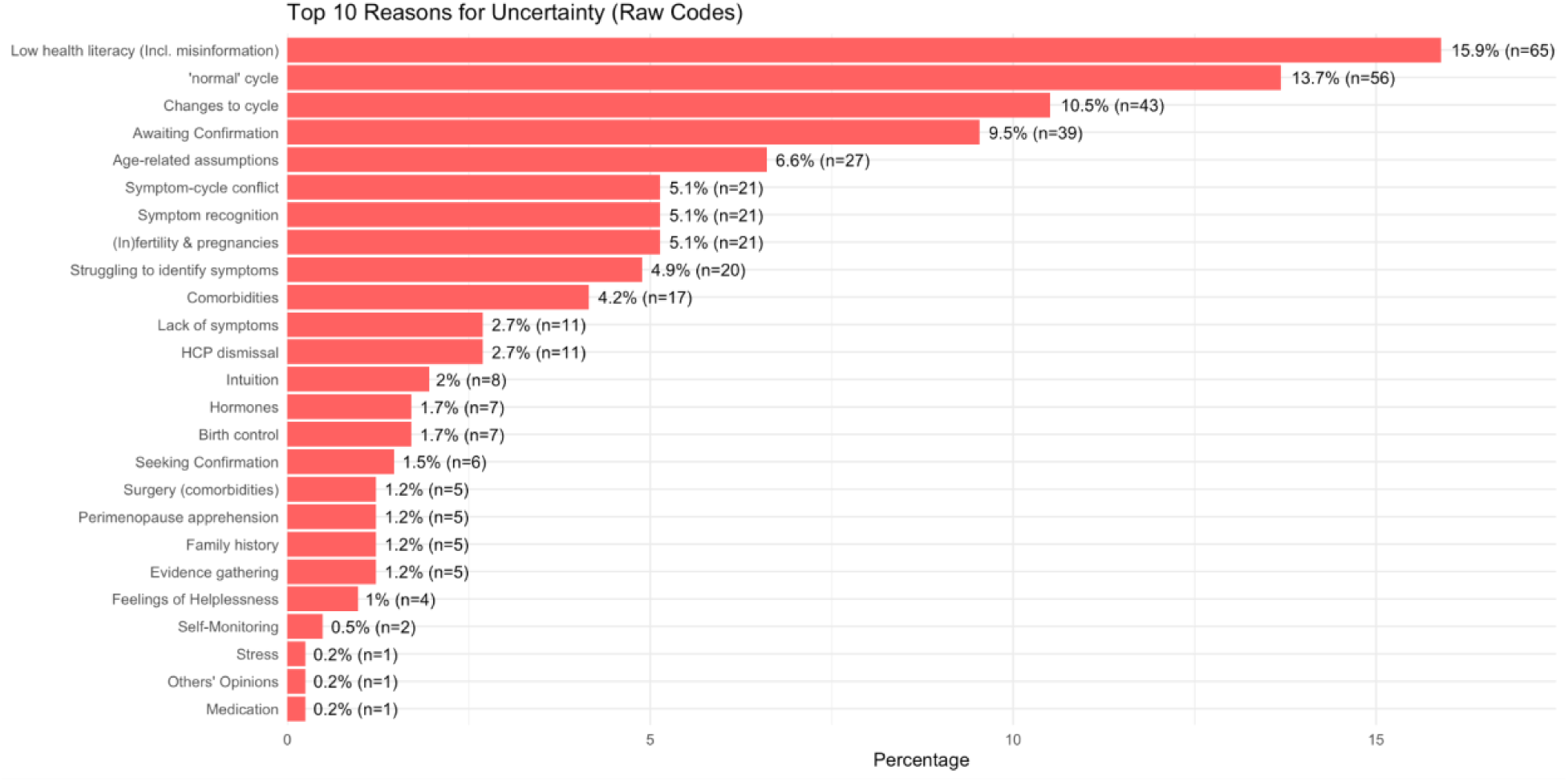
Frequency of raw content-analysis codes among women reporting uncertainty about reproductive stage (n = 409)

